# Quantitative SARS-CoV-2 antibody screening of healthcare workers in the southern part of Kyoto city during the COVID-19 peri-pandemic period

**DOI:** 10.1101/2020.05.12.20098962

**Authors:** Kohei Fujita, Shinpei Kada, Osamu Kanai, Hiroaki Hata, Takao Odagaki, Noriko Satoh-Asahara, Tetsuya Tagami, Akihiro Yasoda

## Abstract

**Background:** The coronavirus disease-2019 (COVID-19) pandemic is associated with a heavy burden on the mental and physical health of patients, regional healthcare resources, and global economic activity. While our understanding of the incidence and case-fatality rates increases, data on seroprevalence of antibodies against the severe acute respiratory syndrome-coronavirus-2 (SARS-CoV-2) in healthcare workers during the peri-pandemic period is insufficient. This study quantitatively evaluated seroprevalence of SARS-CoV-2 antibody in healthcare workers in the southern part of Kyoto city, Japan.

**Methods:** We prospectively recruited healthcare workers from a single hospital between April 10 and April 20, 2020. We collected serum samples from these participants and quantitatively evaluated SARS-CoV-2 IgG antibody levels by enzyme-linked immunosorbent assay.

**Results:** Five (5.4%), 15 (16.3%), and 72 (78.3%) participants showed positive, borderline, and negative serum SARS-CoV-2 IgG antibody status, respectively. We found the mean titer associated with each antibody status (overall, positive, borderline, and negative) was clearly differentiated. Participants working at the otolaryngology department and/or having a history of seasonal common cold symptoms had a significantly higher titer of SARS-CoV-2 IgG antibody (p=0.046, p=0.046, respectively).

**Conclusions:** Five (5.4%) and 15 (16.3%) participants tested positive and borderline, respectively, for SARS-CoV-2 IgG antibody during the COVID-19 peri-pandemic period. These rates were higher than expected based on government situation reports. The present findings suggest that COVID-19 was already spread in the southern part of Kyoto city at the early stage of pandemic.

## Introduction

Coronavirus disease-2019 (COVID-19) is caused by the severe acute respiratory syndrome coronavirus-2 (SARS-CoV-2). COVID-19 was first reported in Wuhan, China, in December 2019; the outbreak was subsequently declared a pandemic by the World Health Organization (WHO) on March 11, 2020 (1). The disease course varies from mild and self-limiting upper respiratory infection symptoms to severe respiratory failure, which might require respiratory support (2, 3). By mid-March 2020, the centers of pandemic had been located in China, the United States, and several European countries. In Japan, the government announced a state of emergency on April 4, 2020. As of the end of April 2020, over 200,000 people worldwide have died of COVID-19 (1, 4). COVID-19 is associated with a heavy burden on mental and physical health of patients, regional healthcare resources, and global economic activity. Effective policies to deal with the pandemic are required and they should be founded on reliable epidemiological data. The diagnosis of COVID-19 is based on viral nucleic acid detection using reverse-transcription polymerase chain reaction (RT-PCR) assay for SARS-CoV-2. Whereas RT-PCR assay is accurate at detecting an active case of COVID-19, identifying subjects who had recovered from SARS-CoV-2 infection has been challenging. In contrast to tracking active cases, antibody detection can provide information on individual and herd acquired immunity against SARS-CoV-2. Furthermore, an antibody assay can help estimate the number of people within a community who remain potential cases, assisting governments in effective decision-making. To-date, data on seroprevalence of SARS-CoV-2 antibody in healthcare workers worldwide is limited. During the peri-pandemic period, we quantitatively evaluated seroprevalence of SARS-CoV-2 antibody in healthcare workers in the southern part of Kyoto city, which is an area famous for its heritage status and a popular tourist destination.

## Participants and methods

### Participants

This study was conducted at the National Hospital Organization Kyoto Medical Center (600 beds), located in southern Kyoto, Japan. In response to the pandemic, our hospital formed an infectious disease department dedicated to COVID-19, involving medical staff such as internal medicine physicians, chest physicians, general and thoracic surgeons, cardiologists, nephrologists, otolaryngologist, and emergency physicians. We prospectively recruited medical doctors, nurses, and ward clerks employed at our hospital between April 10 and April 20, 2020. All participants were asymptomatic and belonged to any of following sections: infectious disease, respiratory medicine, otolaryngology, or an emergency medicine department. We selected healthcare workers from these departments as they were more likely to treat suspected COVID-19 cases, of which they might not have been aware. Additionally, we collected the following questionnaire-based data: history of seasonal common cold from winter 2019 to early spring 2020 and of regular contact with children aged under 12 years. These questionnaires were created based on previous studies on behavior patterns during the H10N8 avian influenza outbreak (5).

### ELISA assay

We collected 6 ml of blood from each participant between April 10 and April 20, 2020. After extracting serum, we stocked them at −80°C deep freezer. We evaluated the presence of serum IgG antibody against SARS-CoV-2 by enzyme-linked immunosorbent (ELISA) assay, using the COVID-19 IgG ELISA kits (DRG international, Inc. Springfield, NJ, USA), as instructed by the manufacturer. Briefly, 1:100 diluted human serum samples were placed onto the 96-well microplate (coated with SARS-CoV-2 recombinant full-length nucleocapsid protein) and then incubated for 30 minutes at room temperature (20-25 °C). After washing, 100 μl HRP-labeled anti-IgG tracer antibody was added into the wells and then the samples were incubated for 30 minutes at room temperature (20-25 °C). Following the second wash cycle, 100 μl substrate was added into the wells and the samples were incubated for 20 minutes at room temperature (20-25 °C). At last, stop solution was added into the wells to terminate the reaction. The optical density of each well was determined by a microplate reader set to 450 nm within 10 minutes. For detection of IgG, the cut off value was modified by using an inner negative control. We interpreted the results as “positive,” “borderline,” and “negative,” following manufacturer’s instructions.

### Statistical analysis

The data were analyzed using JMP version 14.0.0 (SAS institute Inc. Cary, NC). The Fisher’s exact test was used to compare proportions among occupations, wards, questionnaire and SARS-CoV-2 IgG antibody status. Wilcoxon rank sum tests or Kruskal-Wallis tests, as appropriate, were used to compare the titer of SARS-CoV-2 IgG antibody between groups. P-values <0.05 were considered statistically significant.

### Ethical approval

This study was approved by the relevant institutional review boards (approved number 20-009). We obtained written consent form all study participants.

## Results

A total of 92 healthcare workers were recruited for this study. Medical doctors, nurses, and medical clerks constituted 42 (45.7%), 48 (52.2%), and 2 (2.2%) participants, respectively. Of 92 participants, 59 (64.1%) were women; the majority of participants were in their twenties and thirties. The otolaryngology department was the common place of work among the participants, followed by respiratory and emergency medicine departments. There were 47 (51.1%) participants with history of seasonal common cold symptoms from winter 2019 to early spring 2020. There were 19 (20.7%) participants with history of regular contact with children aged under 12 years (Table 1).

**Table 1.** Clinical and demographic characteristics of participating healthcare workers

|  | n=92 |
| --- | --- |
| Gender (female) | 59 (64.1) |
| Age group (years) |  |
| 20s | 30 (32.6) |
| 30s | 29 (31.5) |
| 40s | 21 (22.8) |
| ≥50s | 12 (13.0) |
| Occupation |  |
| Medical doctor | 42 (45.7) |
| Nurses | 48 (52.2) |
| Medical clerk | 2 (2.2) |
| Department |  |
| Department of Infectious Diseases | 18 (19.6) |
| Respiratory Medicine Ward | 22 (23.9) |
| Otolaryngology Ward | 30 (32.6) |
| Emergency Medicine Ward | 22 (23.9) |
| Questionnaire |  |
| *History of seasonal common cold symptoms | 47 (51.1) |
| *History of regular contact with children | 19 (20.7) |
| **History of exposure to a viral infection | 84 (91.3) |
| SARS-CoV-2 antibody status |  |
| Positive | 5 (5.4) |
| Borderline | 15 (16.3) |
| Negative | 72 (78.3) |

Data are shown as counts (%). SARS-CoV-2, severe acute respiratory syndrome coronavirus-2
\*Covered period from winter 2019 to early spring 2020, including history of regular contact with
children aged under 12 years.
\*\*Participants considered exposed to viral infection were defined as those with own history of seasonal common cold symptoms and/or examining outpatients with common cold symptoms.

### Seroprevalence of antibody against SARS-CoV-2

A total 92 serum samples collected between April 10 and April 20, 2020 were tested for antibody against SARS-CoV-2. Of 92 participants, 5 (5.4%), 15 (16.3%), and 72 (78.3%) showed positive, borderline, and negative results of SARS-CoV-2 IgG antibody test, respectively (Table 1). There were no significant differences in antibody status between professional groups (Table 2). There were two and three participants with positive antibody status in the respiratory disease and otolaryngology departments, respectively. The highest proportion of participants with positive and borderline SARS-CoV-2 IgG antibody status was working at the otolaryngology department. In contrast, the lowest proportion of such participants was working at the emergency medicine department (Table 3). Participants with a history of seasonal common cold from winter 2019 to early spring 2020 showed higher rate of positive SARS-CoV-2 IgG antibody testing than did participants without such history (p=0.046). History of regular contact with children or of exposure to a viral infection did not affect the seroprevalence of SARS-CoV-2 IgG antibody (Table 4).

**Table 2.** SARS-CoV-2 IgG antibody seroprevalence among healthcare workers by occupation

| Occupation | n | IgG against SARS-CoV-2 |  |  |  |  |  | P-value |
| --- | --- | --- | --- | --- | --- | --- | --- | --- |
|  |  | Positive |  | Borderline |  | Negative |  |  |
| Medical doctor | 42 | 2 | (4.7%) | 6 | (14.0%) | 34 | (76.2%) | 0.9236 |
| Nurse and Medical clerk | 50 | 3 | (6.0%) | 9 | (18.0%) | 38 | (76.0%) |  |

Data are shown as counts (%). P-value was estimated using the Fisher's exact test. $P < 0.05$ was
considered statistically significant. SARS-CoV-2, severe acute respiratory syndrome coronavirus-2

**Table 3.** SARS-CoV-2 IgG antibody seroprevalence among healthcare workers by department

|  | n | IgG against SARS-CoV-2 |  |  |  |  |  | P-value |
| --- | --- | --- | --- | --- | --- | --- | --- | --- |
|  |  | Positive |  | Borderline |  | Negative |  |  |
| Department of Infectious Diseases | 18 | 0 | (0.0%) | 3 | (16.7%) | 15 | (83.3%) | 0.2102 |
| Respiratory Diseases Ward | 22 | 2 | (9.1%) | 4 | (18.2%) | 16 | (72.7%) |  |
| Otolaryngology Ward | 30 | 3 | (10.0%) | 7 | (23.3%) | 20 | (66.7%) |  |
| Emergency Medicine Ward | 22 | 0 | (0.0%) | 1 | (4.6%) | 21 | (95.5%) |  |

Data are shown as counts (%). P-value was estimated using the Fisher's exact test. $P < 0.05$ was considered statistically significant. SARS-CoV-2,
severe acute respiratory syndrome coronavirus-2

**Table 4.** Seroprevalence of SARS-CoV-2 IgG antibody by exposure status determined by a questionnaire

|  | n | IgG against SARS-CoV-2 |  |  |  |  |  | P-value |
| --- | --- | --- | --- | --- | --- | --- | --- | --- |
|  |  | Positive |  | Borderline |  | Negative |  |  |
| *History of seasonal common cold symptoms | 47 | 5 | (10.6%) | 9 | (19.2%) | 33 | (70.2%) | 0.0458 |
| *History of regular contact with children | 19 | 1 | (5.3%) | 1 | (5.3%) | 17 | (89.5%) | 0.3294 |
| **History of exposure to a viral infection | 84 | 5 | (6.0%) | 14 | (16.7%) | 65 | (77.4%) | >0.99 |

Data are shown as counts (%). P-value was estimated using the Fisher's exact test. P <0.05 was considered statistically significant. SARS-CoV-2, severe acute respiratory syndrome coronavirus-2
\*Covered period from winter 2019 to early spring 2020, including history of regular contact with children aged under 12 years.
\*\*Participants considered exposed to viral infection were defined as those with own history of seasonal common cold symptoms and/or examining outpatients with common cold symptoms.

### Serum SARS-CoV-2 IgG antibody titer

Mean antibody titer of all participants was 0.120±0.0372 (Figure 1A). Mean titer of the antibody positive, borderline, and negative group was 0.219±0.051, 0.161±0.0101, and 0.105±0.018, respectively (Figure 1B). Mean antibody titers stratified by occupation and department are shown in Figure 2A and 2B. There were no significant differences in mean antibody titer between doctors, nurses, and medical clerks (Figure 2A, 0.119±0.0326 and 0.121±0.0058, p=0.994). The mean antibody titer among workers at the otolaryngology department was significantly higher than that among workers elsewhere (Figure 2B, 0.112±0.029, 0.121±0.043, 0.134±0.043 and 0.11±0.018, p=0.046). Participants with a history of seasonal common cold symptoms had a significantly higher titer of SARS-CoV-2 IgG antibody than those without such history (Figure 3A, 0.13±0.044 and 0.11±0.026, p=0.046). There were no significant differences in the mean antibody titer between participants with and without history of regular contact with children or with history of exposure to a viral infection (Figure 3B, p=0.304, Figure 3C, p=0.418).

**Figure 1.**
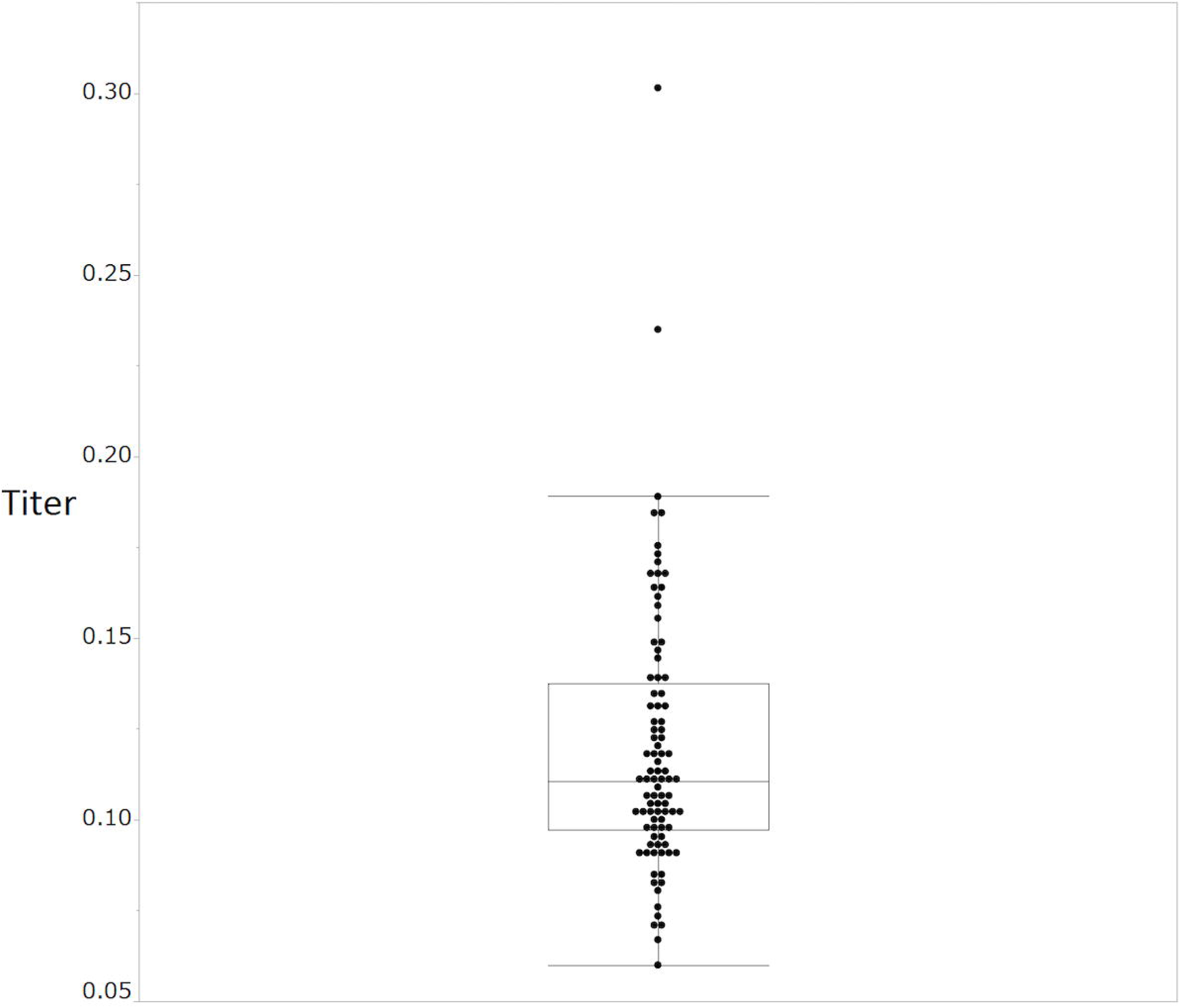
SARS-CoV-2 IgG antibody titers of the participants. (A) All participants (black), (B) stratified by positive (red), borderline (blue), and negative (green) status. Boxes correspond to the interquartile range of values for each group; error bars show the 90th percentile range.

**Figure 2.**
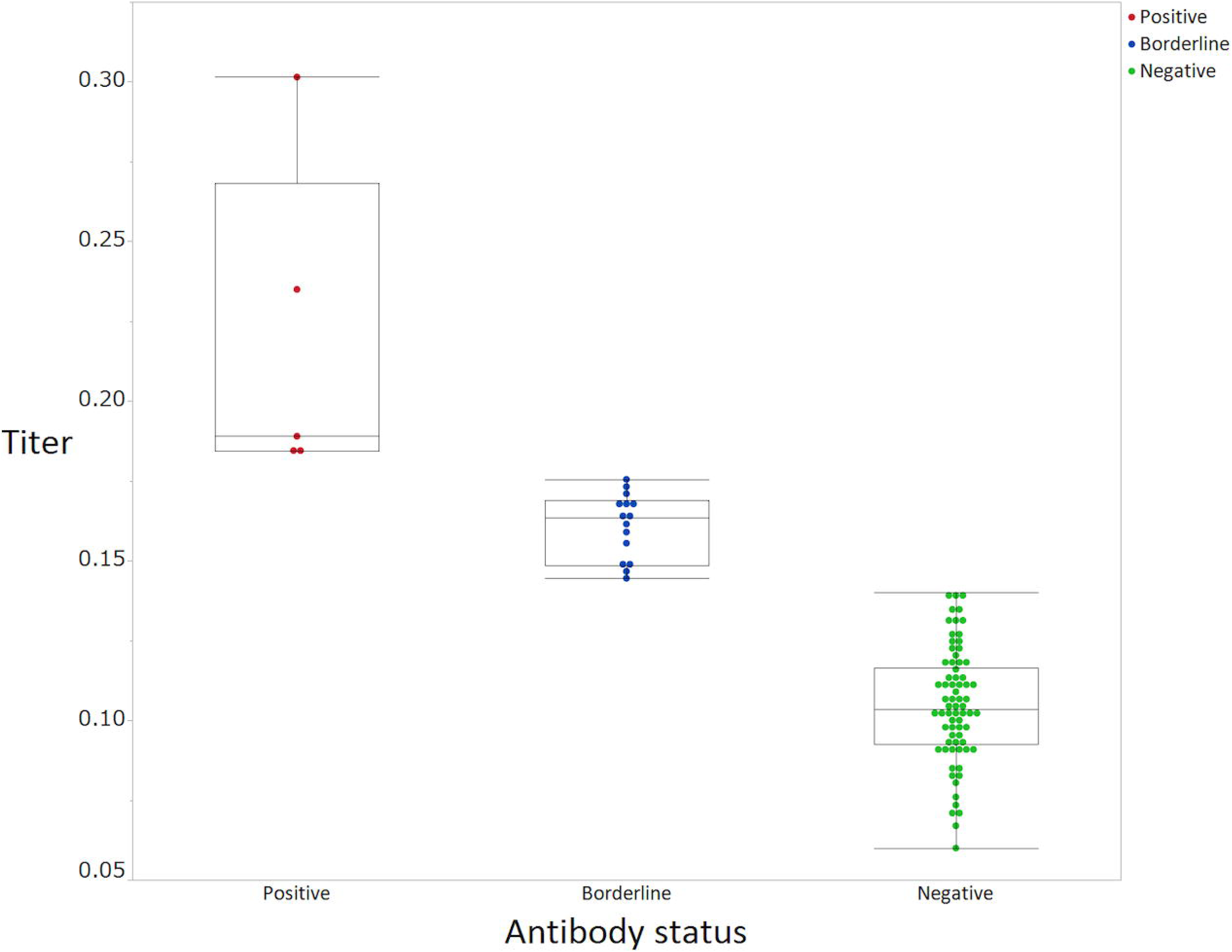
SARS-CoV-2 IgG antibody titers stratified by occupation (A) and department (B). Wilcoxon rank sum test (A) or Kruskal-Wallis test (B) was used to compare the titer of SARS-CoV-2 IgG antibody level between the groups. P-values <0.05 were considered statistically significant.

**Figure 3.**
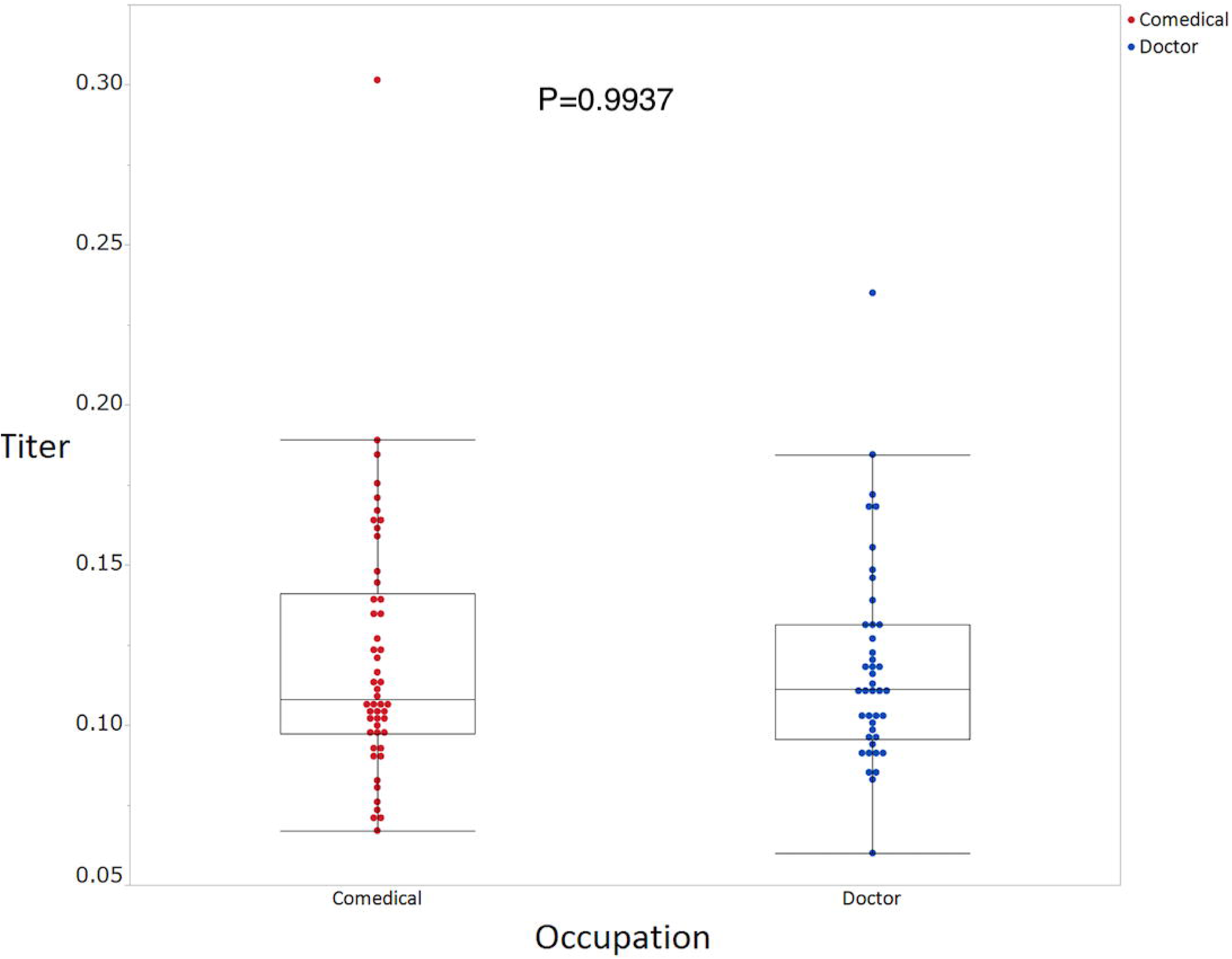
SARS-CoV-2 IgG antibody titers stratified by exposure status determined by questionnaires concerning behavior patterns (A-C). Wilcoxon rank sum tests were used to compare the titer of SARS-CoV-2 IgG antibody levels between the groups. P-value <0.05 were considered statistically significant.

## Discussion

In this study, 5 (5.4%) and 15 (16.3%) healthcare workers, respectively, were “positive” and “borderline” for the presence of SARS-CoV-2 IgG antibody. Mean antibody titer among the borderline group was clearly distinct from that of the negative group. Participants with borderline antibody results might have been latently sensitized by patients with COVID-19. As our hospital accepted patients with confirmed COVID-19 after April 15, 2020, the antibody status of present study participants might reflect community-acquired immunity, resulting from unconscious exposure in daily medical practice.

The mean antibody titer was significantly higher among workers at the otolaryngology department than among those working elsewhere. This suggests that healthcare workers within the otolaryngology department were more likely to be exposed to SARS-CoV-2 than were their counterparts at other departments.

According to the official statements of Kyoto city, confirmed incidence and fatality associated with COVID-19 in Kyoto city at the end of April 2020 were 215 cases and 11 cases, respectively (6). However, antibody seroprevalence observed in the present study was much higher than that expected based on the data from government reports. Furthermore, the number of participants with borderline antibody status in our study was nearly three times that of those with positive antibody status. Recently, several studies have revealed that population-wide seroprevalence of SARS-CoV-2 antibodies was higher than expected based on the number of confirmed cases (7, 8). For example, in Kobe city, Japan, 3.3% of outpatients tested positive for SARS-CoV-2 IgG antibody (8). By using quantitative methods, our results revealed there were more than tripled people sensitized with SARS-CoV-2 on the flip side of people with positive SARS-CoV-2 IgG antibody. Given that pathogenicity of SARS-CoV-2 seems to be similar to SARS-CoV, most patients infected SARS-CoV-2 will express specific IgG antibody within a week to three months after infection.(9) In light of this timeline of seroconversion, the participants with positive and borderline antibody status in the present study were likely exposed to SARS-CoV-2 between December 2019 and March 2020. These findings suggest that COVID-19 was already present in Kyoto at the early stages of pandemic. The period between December and March is a time of heightened tourist activity in Kyoto, in particular, involving tourists from China and Taiwan who celebrate the Chinese New Year spring festival. After the spring festival, on March 5, 2020, the Japanese government implemented a strict ban on travelers arriving from China.

According to epidemiological data provided by the WHO (1) and Johns Hopkins University (4), incidence and case-fatality rates in major European countries (Germany, United Kingdom, France, Italy, and Spain) and the United States are much higher than those in major Asian countries (China, Japan, South Korea, and Taiwan). There are several possible explanations of this phenomenon. Differences in lifestyle and behavioral habits between Western and Asian populations might explain some of the variability in rates. Some studies have shown a correlation between universal BCG vaccination policy, and morbidity and mortality associated with COVID-19 (10, 11). Although this hypothesis has resulted in clinical trials to evaluate the efficacy of BCG vaccination against COVID-19 (NCT04327206 and NCT04362124), restricted basic and clinical evidence makes this association difficult to evaluate. Meanwhile, other authors have suggested that differences in viral genotypes and virulence may affect morbidity and mortality associated with COVID-19; however, this explanation is insufficient. The National Institute of Infectious Diseases of Japan has revealed that the first wave of COVID-19 pandemic emerged from Wuhan, China, and it flattened toward the end of March; however, the second wave emerged from European countries, spreading across the country after the end of March (12). Nevertheless, even during the second wave, Japan retained its much lower morbidity and mortality rates than those of Western countries, as reported at the end of April 2020 (13). Kamikubo and colleague have hypothesized that the pre-pandemic spread of low-virulence type of SARS-CoV-2 and subsequent exposure to mild-virulence type of SARS-CoV-2 induced herd immunity that reduced the severity of high virulence type of SARS-CoV-2 in Japan (14). The findings concerning borderline antibody titer in the present study may support this hypothesis. In the present study, participants with a history of seasonal common cold from winter 2019 to early spring 2020 had a significantly higher SARS-CoV-2 antibody titer than did participants without such history, which might have resulted from pre-pandemic exposure to low- and middle-virulence type of SARS-CoV-2. Further research is required to verify this hypothesis.

There are several limitations to this study. First, as this was a single-center study, selection bias might have affected our findings. Second, the small sample size restricted the statistical power of our analyses. Third, as our participants were recruited from departments where exposure to COVID-19 was more likely, the reported seroprevalence might be an overestimate relative to that of workers at other departments or the general population. Fourth, because the COVID-19 pandemic is an ongoing emerging situation, a significant proportion of available research might be premature; in our discussion, we have referred to such studies.

In conclusion, we have shown relatively high frequency of positive and borderline SARS-CoV-2 antibody status in healthcare workers in the southern part of Kyoto city, an area frequented by tourists. Our results suggest that COVID-19 might have already been present in Kyoto at the early stage of pandemic. Several previous studies have evaluated SARS-CoV-2 antibody profiles in patients with COVID-19 (15-17); however, our study is the first to quantitatively evaluate antibody levels in healthcare workers involved with patients during the COVID-19 peri-pandemic period. Serial evaluation of SARS-CoV-2 IgG antibody will reveal risk factors associated with COID-19 susceptibility and mechanisms of disease spread. Finally, these results should be approached with caution, as there remains a lack of evidence regarding the role of antibodies present after recovery from COVID-19 in developing immunity against subsequent infections.

## Data Availability

All data of this study is available for anyone if our institute IRB allows.

## Author contributions

KF and SK designed of the study and collected blood samples. KF, SK, OK, HH and TO recruited participants and collected data. TT, NSA and AY managed ELISA tests and interpretation of results. KF, SK and OK participated in statistical analysis. KF and SK drafted and revised manuscript. All authors reviewed and revised manuscript.

## Acknowledgement

The authors appreciated the contributions of Takayuki Inoue and Chinami Shiraiwa who conducted the ELISA tests. The authors thank Drs. Koji Hasegawa and Hiroshi Okuno for their advice and support in obtaining an IRB approval. The authors also thank the members of the department of infectious diseases (Drs. Eriko Kashihara, Kosuke Doi, Syuhei Ikeda, Daisuke Hirai, Masayuki Hashimoto and Koichi Seta) and emergency medicine (Drs. Hiroyuki Tanaka, Satoru Beppu and Kei Nishiyama) for their assistance with this study and clinical management of COVID-19.

---

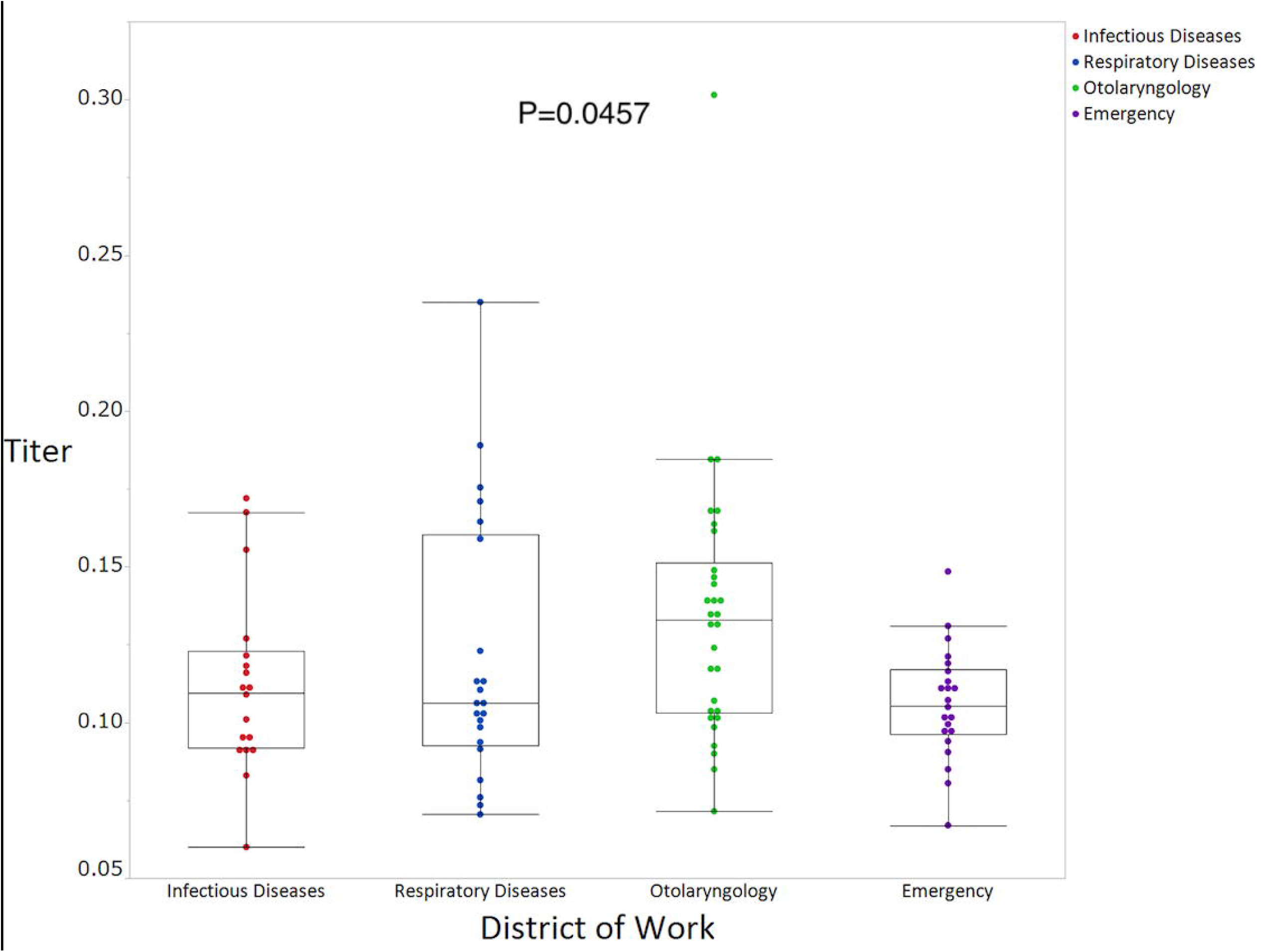

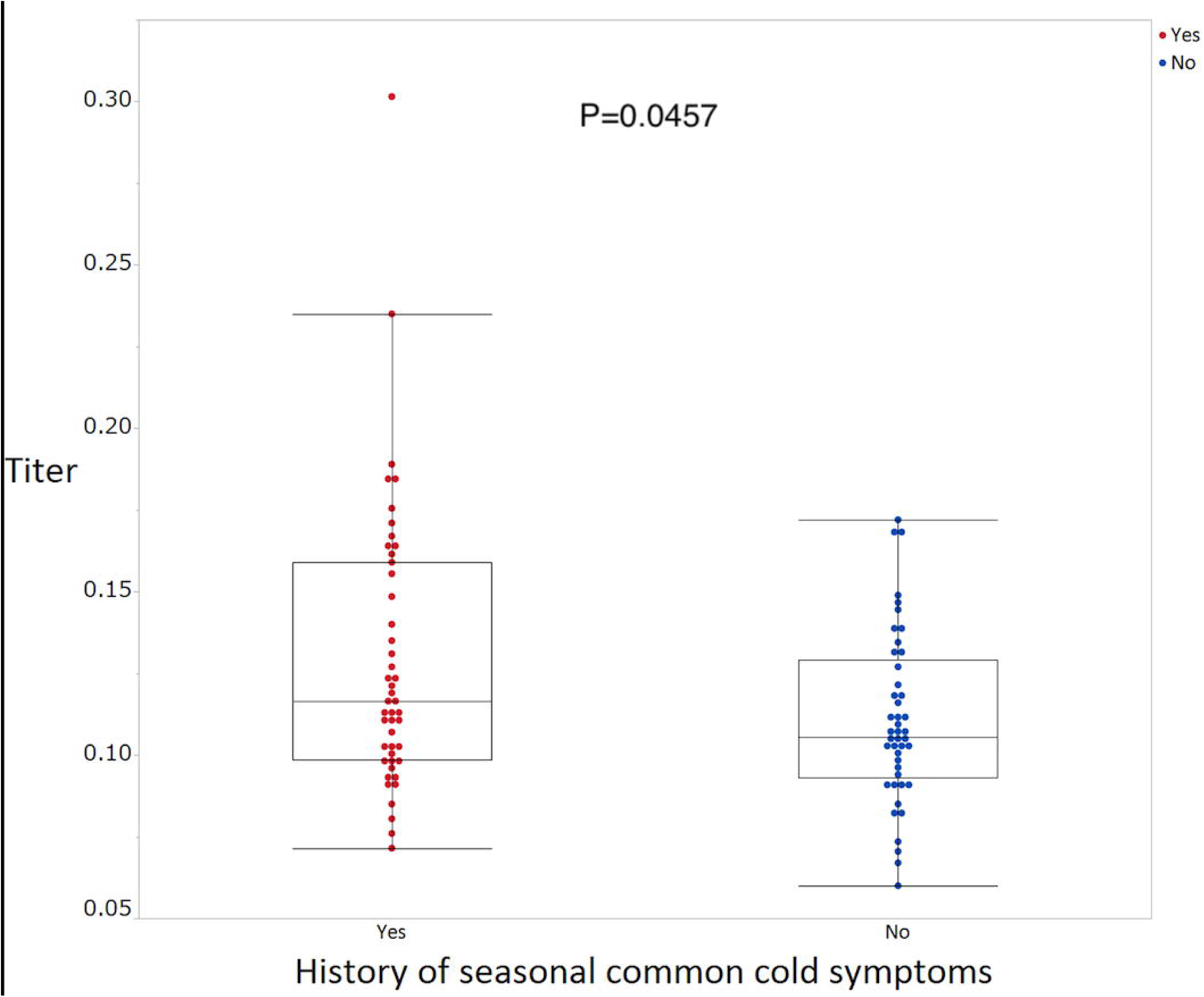

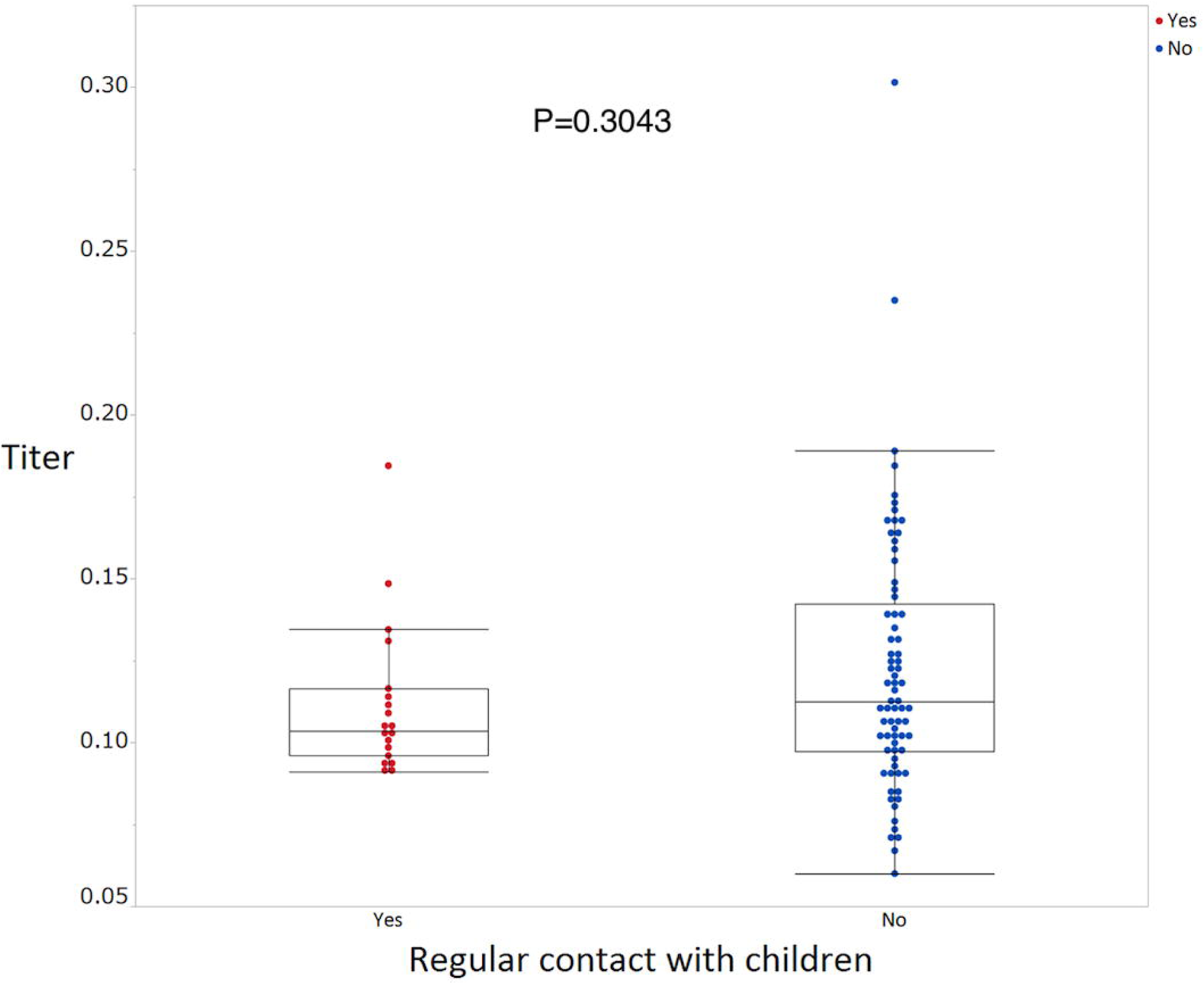

